# Healthcare Worker s (HCWs) Knowledge and Attitudes Related to Pain Management in a Lebanese Tertiary Hospital

**DOI:** 10.1101/2025.10.28.25336335

**Authors:** Maha Kassem, Celine Bakri, Nadine Zeinab, Nadine Hindi, Serene Abou Samra, Zeina Jalabi, Abdelkader Hammoud

## Abstract

**OBJECTIVE:** This study aims to evaluate the knowledge and attitudes of healthcare workers (HCWs) regarding pain assessment and management at a tertiary medical centre in Beirut, Lebanon as well as to identify areas for improvement.

**DESIGN:** A cross-sectional study was conducted with Healthcare Workers using the “Knowledge and Attitudes Survey Regarding Pain” (KASRP), a validated practice-based questionnaire. This specific tool was used to assess the participants’ knowledge and attitudes related to pain management. Descriptive statistics were presented as frequencies, percentages, means, and standard deviations. Independent samples t-tests or ANOVA tests were used to compare mean knowledge scores between groups. A p-value of < 0.05 was considered statistically significant.

**SETTING:** The study was conducted over a period of 1 month at a private tertiary hospital in Beirut, Lebanon.

**PARTICIPANTS:** The study included physicians, nurses, and pharmacists working in different departments within the hospital: Internal Medicine, Pharmacy, Nursing, and Quality Management.

**RESULTS:** A total of 161 HCWs participated in the study (41 physicians, 100 nurses, and 20 pharmacists). The overall mean KASRP score obtained across all participants was 13.65 (SD = 9.93) for correct responses, indicating a moderate level of knowledge. Only 9 participants (5.6%) scored ≥80%, which is considered as a satisfactory level of knowledge and attitude: 4 doctors (9.8%), 5 nurses (5%), and no pharmacists. Items of the KASPR score were categorized into 5 domains: assessment, medication, addiction, spiritual considerations and interventions. Variables potentially affecting knowledge and attitude towards pain management were analyzed: gender, age, profession, prior training on pain management, and years of experience. There was no statistical significance in the mean score of correct answers based on gender, prior training, or years of experience (p >0.05). However, age and profession showed a statistical significance of impact on the mean score of correct answers among HCWs (P <0.05), suggesting that these factors may influence pain management understanding among HCWs

**CONCLUSION:** The findings highlight gaps in knowledge and attitudes toward pain management among HCWs. Structured education and training programs on pain management are deemed necessary to enhance the knowledge and attitudes of HCWs for better patient outcomes.

**WHAT IS ALREADY KNOWN ON THIS TOPIC:** Pain is an undertreated clinical symptom and health burden affecting both hospitalized and non-hospitalized patients. Although pain has been designated as the 5th vital sign to ensure pain assessment and management, many patients continue to have uncontrolled or inadequate management of pain (1)(2). This negatively impacts the patients’ quality of life and overall wellbeing (2). A lack of adequate knowledge among HCWs remains a significant barrier to effective pain management (12), with misconceptions about pain evaluation; incorrect beliefs that patients overstate their pain; fears about drug tolerance and addiction; a lack of understanding of analgesic pharmacology; and difficulties in assessing pain in children (13).

**WHAT THIS STUDY ADDS:** To date, there are no published studies that assessed the knowledge and attitudes of physicians, nurses, and pharmacists toward pain management in Lebanon. Conducting such research is essential to identify discipline-specific gaps and inform strategies to improve pain management knowledge and clinical practice among physicians, nurses, and pharmacists alike.

## Introduction

Pain remains a global health concern that affects individuals of all ages, from different backgrounds, and with different health issues. Despite international guidelines and many interventions to reduce myths and increase awareness among the healthcare professionals and public sector, pain continues to be inadequately controlled or ineffectively managed (3). It is a multidimensional physiological and psychological phenomenon, acute or chronic, that is inherently subjective and can be defined as an unpleasant sensory and emotional experience related to actual or potential tissue damage (4)(5). Pain has been designated as the 5th vital sign to emphasize the importance of its assessment and management (6). Untreated or inadequately treated pain can reduce a person’s quality of life (QoL) compared to the general population and those with other limiting conditions, highlighting the need for accurate assessment, and efficient and effective pain management (7)(8). It can also lead to adverse physiological and psychological outcomes for both patients and their families, while also increasing the burden on healthcare systems through extended hospital stays, higher readmission rates, and increased healthcare costs (9).

Effective pain management requires an interdisciplinary approach by various healthcare workers (HCWs), including nurses, physicians, and pharmacists with a strong foundation of knowledge and the right attitude. Such knowledge is believed to affect the attitude of the HCWs in managing pain (10)(11). However, a lack of adequate knowledge among HCWs remains a significant barrier to effective pain management (12), with misconceptions about pain evaluation; incorrect beliefs that patients overstate their pain; fears about drug tolerance and addiction; a lack of understanding of analgesic pharmacology; and difficulties in assessing pain in children (13).

Similar key clinical barriers to effective pain management include insufficient knowledge (14), opioid phobia related to addiction and respiratory depression (15), poor interdisciplinary communication (16), reluctance among medical teams to prescribe opioids (17), delays in opioid storage protocols (18), and inadequate pain assessment, lack of protocols, and nursing workload (19).

Several studies in the middle east region have shown an array of deficit in knowledge and attitudes of HCWs toward pain management (20)(21)(22).

In Lebanon, 2 studies addressed the knowledge and attitudes of physicians and nurses, separately. Nasser et al. evaluated physicians’ self-assessed pain management skills and identified barriers to effective pain control and identified fear of adverse events, particularly opioid side effects as barriers and a lack of adequate knowledge about pain management (2). Abdul Rahman, Abu-Saad Huijer, and Noureddine identified a lack of knowledge with only 3.4 % of the nurses achieving a score of equal to or greater than 80%, the required score for effective pain management (23).

To date, there are no published studies that assessed the knowledge and attitudes of physicians, nurses, and pharmacists toward pain management in Lebanon. Conducting such research is essential to identify discipline-specific gaps and inform strategies to improve pain management knowledge and clinical practice among physicians, nurses, and pharmacists alike.

Accordingly, our study aims to (1) evaluate the knowledge and attitudes of HCWs regarding pain assessment and management at a tertiary medical center in Beirut, Lebanon; and (2) identify areas of deficiency for targeted improvement initiatives.

## Methods

### Data source and study population

The “Knowledge and Attitudes Survey Regarding Pain” (KASRP), a validated practice-based questionnaire, originally developed by Ferrell and McCaffery in 1987 and revised in 2014 was used to assess the participants’ knowledge and attitudes related to pain management in its English form, since the official language used in the hospital was English and all participants were deemed able to read, comprehend and react to the questionnaire in English language (24). The KASPR questionnaire content is based on the standards of pain management such as the American Pain Society, World Health Organization, and the National Comprehensive Cancer Network pain guidelines. The content validity of the tool was established by the original authors, along with the test-retest reliability which was very good (r > 0.8) when tested on a class of staff nurses (N=60), and the internal consistency reliability was also found to be good (r > 0.70) and its authenticity, both in terms of its appearance and contents, was validated by an impartial panel of experts (24).

As per the creators of the tool, questions may be omitted, so we removed some that were mainly related to pediatric pain, culture, and one of the case study. A score of “1” was provided for each correct response and a “0” for the incorrect response. The total score was the sum of all correctly answered questions. The percentage score was calculated by dividing the number of correct responses by the total number of items in the survey.

The questionnaire was self-administered electronically via an online survey link distributed to eligible participants. The KASRP questionnaire was divided into two parts and included a total of 32 items from the KASRP based on our study objectives: (1) The sociodemographic data: this section collected information on participants’ gender, age group, profession (doctor, nurse, and pharmacist), clinical specialty, years of working experience, and any prior education or training on pain management; and (2) Knowledge and Attitudes Assessment: the second part included the amended KASRP questionnaire with 19 True/False questions, 12 Multiple Choice Questions, and One clinical case study designed to evaluate participants’ knowledge and attitudes regarding various aspects of pain assessment and management. Data collection took place over 1 month in 2025.

HCWs were considered to have adequate knowledge and attitude if the score was 80% and above, a level identified by McCaffery and Robinson (2002). According to Ferrel et al, items should be differentiated with the least correct responses and those with the best scores for better response analysis (24).

### Intervention

#### Study design and setting

A cross-sectional study was conducted over a one-month period, from 23 February to 23 March 2025, at a private tertiary hospital in Beirut, Lebanon, to assess the knowledge and attitudes of HCWs towards pain management.

#### Patient and Public Involvement

The study included HCWs (physicians, nurses, and pharmacists) who were actively employed at the hospital during the study period and consented to participate. The sample size for the study was 161. Inclusion criteria required participants to be licensed HCWs involved in clinical care or pharmaceutical services. Exclusion criteria included HCWs not involved in direct patient care, such as administrative staff and midwives, pediatric nurses and pediatricians. The questionnaire was distributed using an online survey platform to ensure ease of access and confidentiality. Participants received the survey link via their mobile phones and were asked to complete the questionnaire independently without any external assistance to avoid bias. Participation was voluntary, the enrolled staff filled the survey anonymously, and informed consent was obtained electronically before starting the survey questions.

#### Statistical analysis

Collected data were exported from the survey platform and analyzed using IBM SPSS Statistics version 25. Descriptive statistics summarized participants’ demographic characteristics and questionnaire responses, presented as frequencies, percentages, means, and standard deviations where applicable. Inferential statistics were applied to evaluate the association between knowledge scores and demographic variables. Independent samples t-tests or ANOVA tests were used to compare mean knowledge scores across different groups (e.g., profession, specialty, previous education on pain management). One-way ANOVA was used for multi-category variables (e.g., age, profession), and independent samples t-tests were used for binary variables (e.g., gender, training). A p-value of less than 0.05 was considered statistically significant. The clinical case study responses were analyzed separately to assess applied knowledge.

## Results

### Participants’ Characteristics

#### Knowledge and attitude amongst HCWs regarding pain

The overall mean correct KASPR score obtained across all participants was 13.65 (SD = 9.93) for correct responses out of 33 possible points, indicating generally poor knowledge. Only 9 out of 161 participants (5.6%) scored ≥80%: 4 doctors (9.8%), 5 nurses (5%), and 0 pharmacists (Table 2).

A total of 161 healthcare professionals participated in the study. The sample included 83 (51.6%) females and 78 (48.4%) males. Most respondents were between 25 and 45 years old (72.1%), with the largest subgroup aged 36 and 45 years (37.3%). Professionally, nurses made up the majority of the sample (62.1%), followed by doctors (25.5%) and pharmacists (12.4%). In terms of experience, 41% had more than 15 years, while 23.6% had less than 5 years. The demographic characteristics are tabulated in Table 1.

**Table 1.** Demographic Characteristics of Participants (n = 161)

| Characteristic | Category | Frequency | Percentage (%) | P-Value |
| --- | --- | --- | --- | --- |
| **Gender** | Female | 83 | 51.6 | 0.513 |
|  | Male | 78 | 48.4 |  |
| **Age Group** | 20–24 years | 14 | 8.7 | *0.021 |
|  | 25–35 years | 56 | 34.8 |  |
|  | 36–45 years | 60 | 37.3 |  |
|  | 46–55 years | 24 | 14.9 |  |
|  | > 55 years | 7 | 4.3 |  |
| **Profession** | Nurse | 100 | 62.1 | *0.001 |
|  | Doctor | 41 | 25.5 |  |
|  | Pharmacist | 20 | 12.4 |  |
| **Subspecialty** | Anesthesiology | 10 | 6.2 | 0.171 |
|  | General Practice | 14 | 8.7 |  |
|  | Medicine | 18 | 11.2 |  |
|  | ONCOLOGY | 12 | 7.5 |  |
|  | Surgery | 39 | 24.1 |  |
|  | Other | 65 | 40.4 |  |
| **Years of Experience** | < 5 years | 38 | 23.6 | 0.482 |
|  | 5–10 years | 33 | 20.5 |  |
|  | 11–15 years | 24 | 14.9 |  |
|  | > 15 years | 66 | 41.0 |  |

**Table 2.** Percent and count of responders by specialty who have 80% or more as an average correct score.

| Specialty | Frequency | Percent from total by Specialty |
| --- | --- | --- |
| Physicians | 4 | 9.8% |
| Nurses | 5 | 5% |
| Pharmacists | 0 | 0% |

**Table 3.**
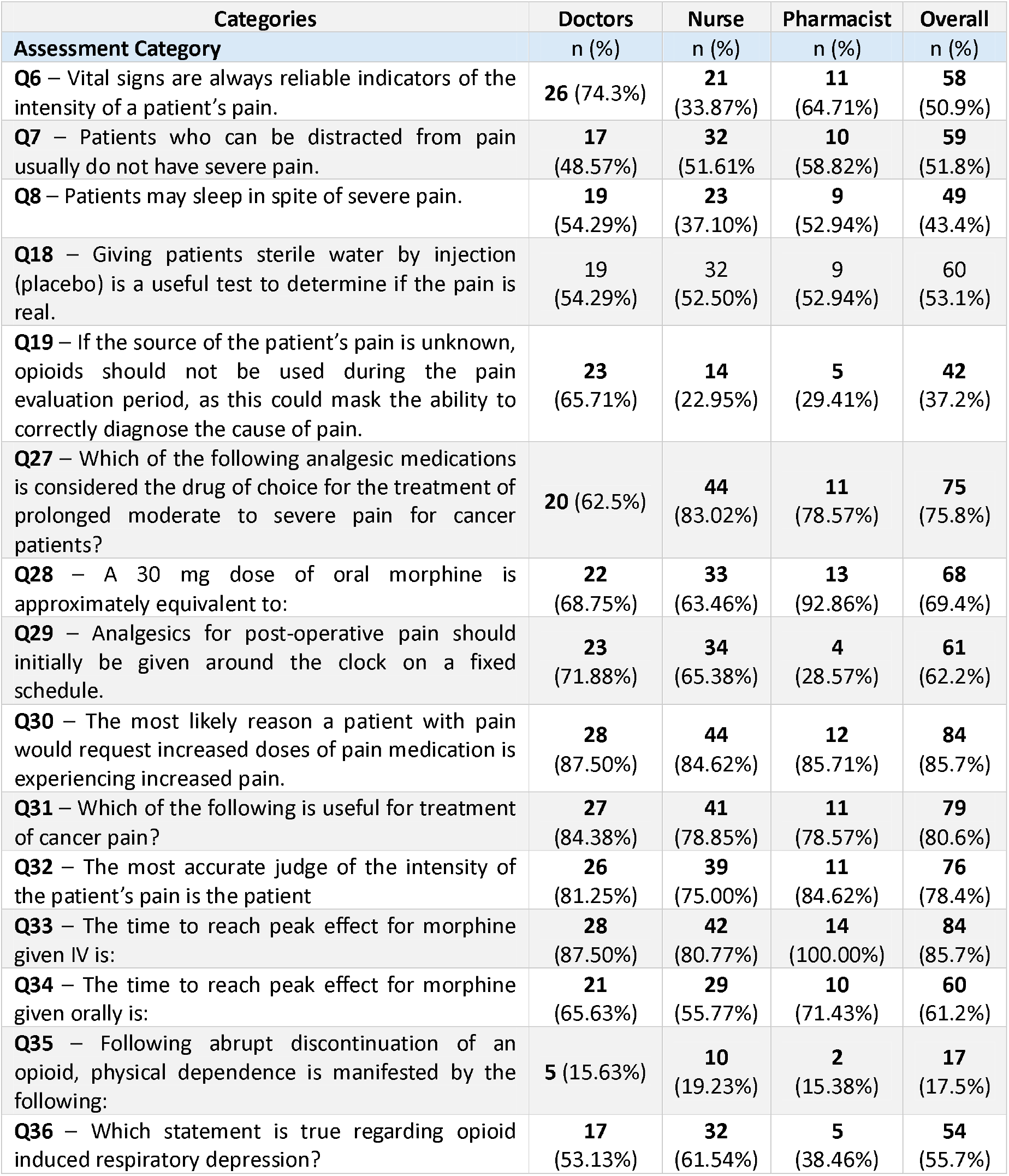
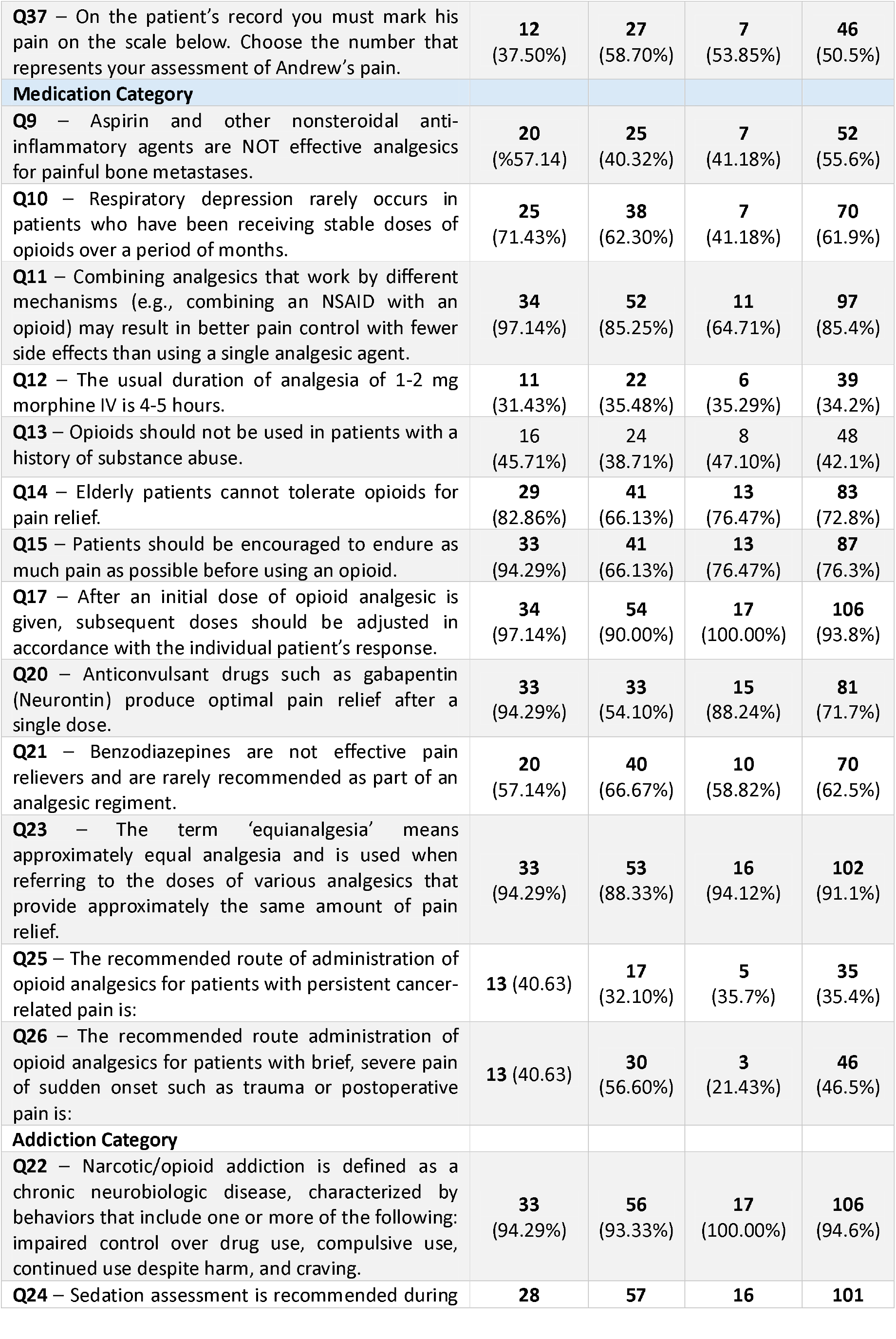
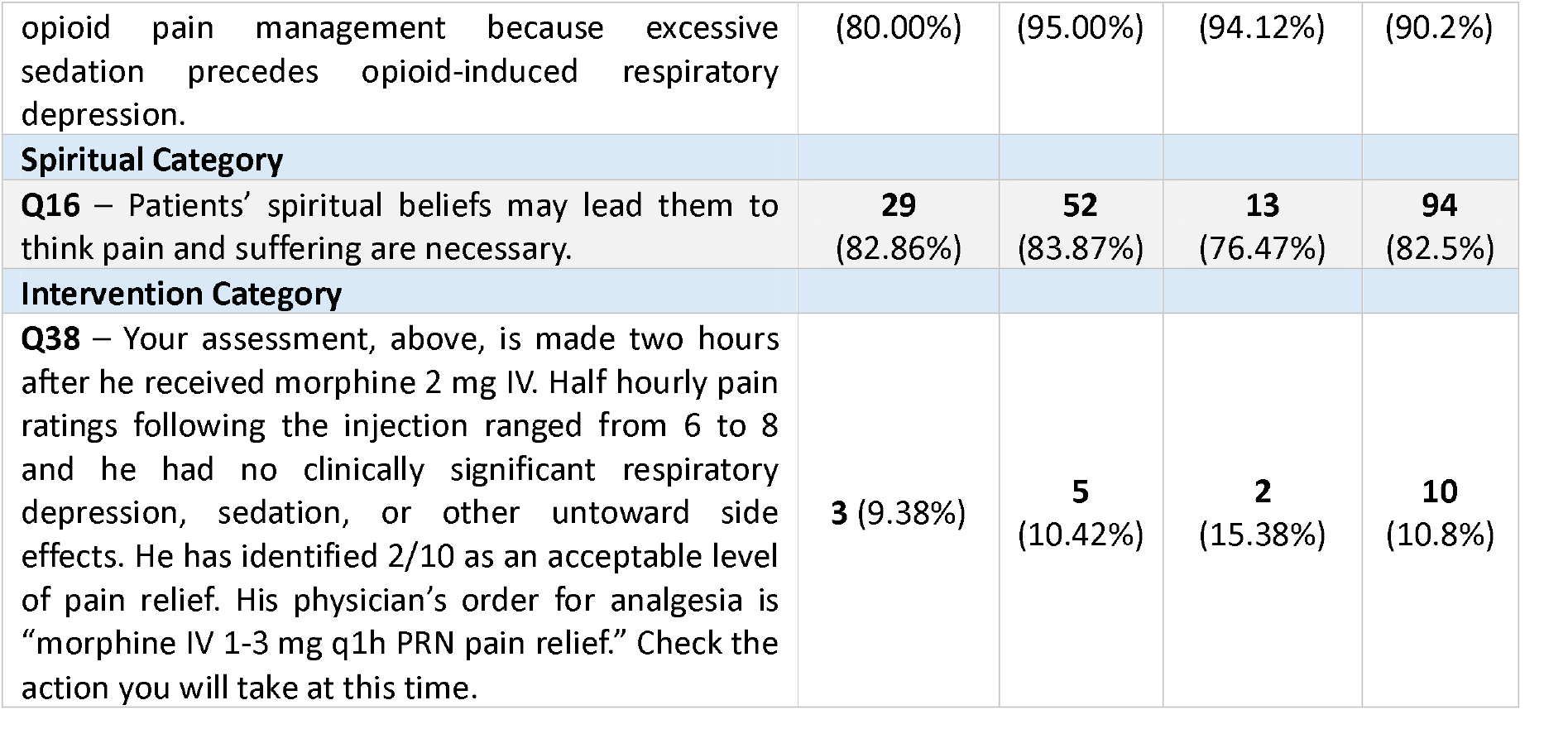
Percentage of Correct Answers by KASRP Item among HCWs based on Question Category.

Factors affecting the knowledge and attitude towards pain management were analyzed: gender, age, profession, training status, and years of experience (Table 4). Gender: Although male participants had a slightly higher mean score (M = 14.74) than females (M = 12.63), this difference was not statistically significant (p = 0.177) based on an independent samples t-test. Age: A one-way ANOVA revealed a statistically significant difference in mean scores among age groups (p = 0.021). Participants aged 46–55 years achieved the highest mean score (M = 18.04), while those aged 25–35 and 36–45 scored the lowest. Profession: There was a highly significant difference in knowledge scores by profession (p = 0.001, one-way ANOVA). Doctors scored the highest (M = 18.05), followed by pharmacists (M = 16.05), with nurses scoring the lowest (M = 11.37). Training in Pain Management: Those who had received formal training in pain management scored higher (M = 15.11) than those who had not (M = 12.88), but this difference was not statistically significant (p = 0.175). Years of Experience: The effect of years of experience on knowledge scores was analyzed using ANOVA and was found to be not statistically significant (p = 0.482).

**Table 4.**
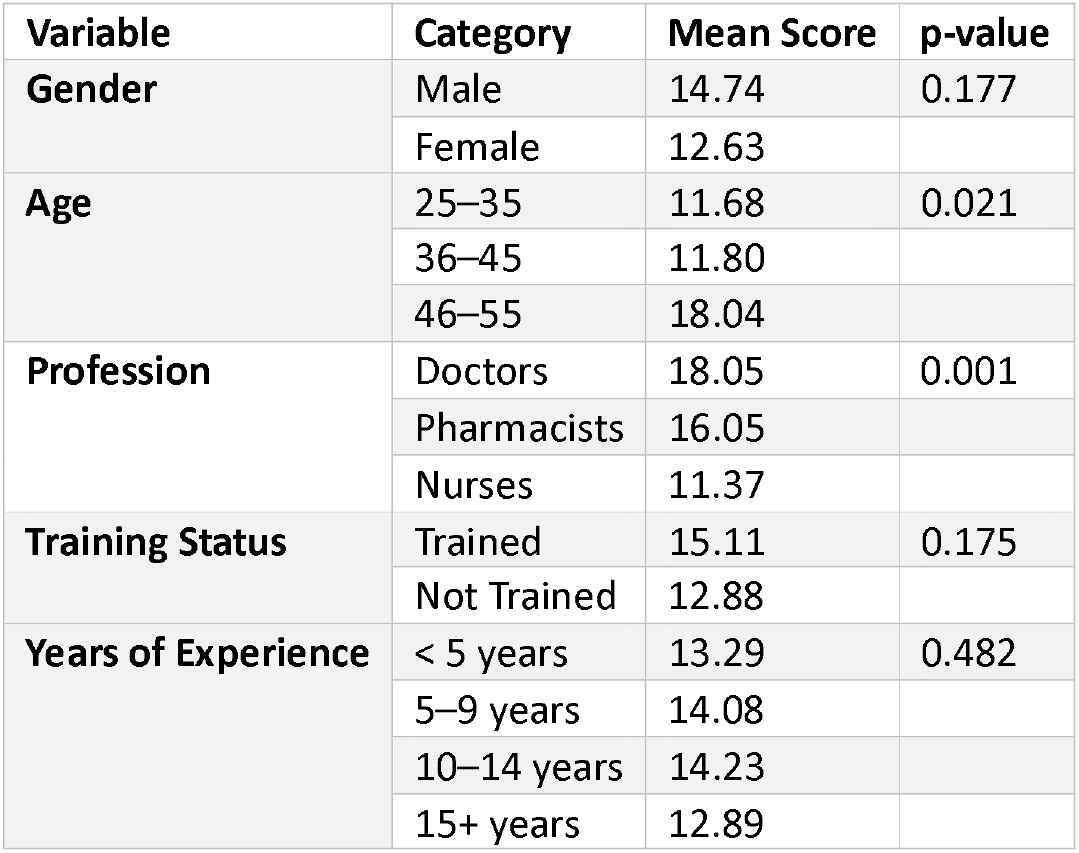
Factors Affecting knowledge on pain management.

| Variable | Category | Mean Score | p-value |
| --- | --- | --- | --- |
| **Gender** | Male | 14.74 | 0.177 |
|  | Female | 12.63 |  |
| **Age** | 25–35 | 11.68 | 0.021 |
|  | 36–45 | 11.80 |  |
|  | 46–55 | 18.04 |  |
| **Profession** | Doctors | 18.05 | 0.001 |
|  | Pharmacists | 16.05 |  |
|  | Nurses | 11.37 |  |
| **Training Status** | Trained | 15.11 | 0.175 |
|  | Not Trained | 12.88 |  |
| **Years of Experience** | < 5 years | 13.29 | 0.482 |
|  | 5–9 years | 14.08 |  |
|  | 10–14 years | 14.23 |  |
|  | 15+ years | 12.89 |  |

## Discussion

The following study investigated the knowledge and attitude of physicians, nurses, and pharmacists toward pain management in a tertiary care hospital in Beirut, Lebanon. The findings from this study demonstrated moderate overall knowledge of pain management among healthcare professionals, with only 5.6% of participants achieving the benchmark score of ≥ 80% (25). Despite some well-answered items, particularly those related to basic opioid pharmacology and assessment, other areas reveal significant knowledge gaps that reflect a comparable pattern in several Middle Eastern and developing countries.

## Main Findings

### Comparison with International Findings

Only 5.6% of the total sample achieved the benchmark score of 80% or higher. Compared to international studies, this rate is substantially lower. In the United States, AlShaer et al. reported that 54% of nurses scored above 80%, while Turkish nurses in Yildirim et al. averaged a 73.3% correct response rate (26)(27). This difference is attributed to better integration of pain education into the nursing curricula and professional training on pain management. In Hong Kong, Lui et al. reported that nurses’ knowledge and attitudes on pain management were significantly influenced by clinical working experience and applied knowledge of pain to daily practice (28). This is comparable to our study where HCW who received formal training in pain management scored higher mean score of correct answers compared to non-trained although statistically non-significant 15.11 vs. 12.88 respectively (P<0.05).

### Comparison with Regional Findings

When considering comparison of our study results with other Middle Eastern countries, our results become more mirrored. In Jordan, Nuseir et al. study results reported that physicians and pharmacist scored 36% and 24% respectively, aligning with our study’s results that nurses had the lowest scores, followed by pharmacists (20). In Saudi Arabia, similarly, Al-Quliti and Alamri reported that only 5.7% of participants scored > 60%, mirroring our study results were only 5.6% of the HCWs achieved 80%, this reflects a regional existing gap at the level of pain management education and practice (21).

This consistency in regional findings across the Middle East region reflects an existing systemic education regional gap despite the differences in institution systems, professions, and languages. The core deficits, as reflected from the percentage of correct answers by the HCWs are diversified and fall into lack of understanding of the opioid pharmacology, fear of addiction, and appropriate assessment.

### Profession-Specific Observations

In our study, with respect to profession-specific observation, physicians scored the highest (mean= 18.05), followed by pharmacists (mean=16.05) and nurses (mean=11.37). This profession-specific observation aligns with findings in Jordan, Saudi Arabia, and Iran, where physicians showed highest scores in comparison to nurses and pharmacist. This can be likely attributed to more medical training. However, only 9.8% of the physicians met the adequacy benchmark ≥80%, reflecting that profession does not necessarily ensure competency in pain management unless adequate education and professional training exists in place. In our study, none of the pharmacists scored ≥80% although pharmacists are positioned to evaluate prescribed pain medications and treatment plan. This can be attributed to limited clinical experience in pain management, lack of adequate training, and underestimation of the role of the pharmacist in appropriate pain management. This contrasts with the Western world where pharmacists are involved in pain stewardship programs and pain management.

### Impact of Training and Experience

HCWs who received pain management training scored slightly higher than those who did not, mean of 15.11 vs. 12.88, although not statistically significant (p=0.175). Similarly, this aligns with the findings of Al-Quliti & Alamri and Nuseir et al. emphasizing the importance of education, training, and curricula integration of pain management (21)(20). There was no statistically significant difference in mean correct scores observed based on gender or years of experience, indicating that experience alone does not replace training and education. The age revealed a statistical significance (P<0.05) in the mean scores among age groups, with age groups 46-55 scoring the highest mean score of (18.04) among the other age group of 25-35 (11.68) and 36-45 (11.08) respectively.

### Limitations

This research should be interpreted in light of a number of limitations. Being conducted in a single tertiary hospital in Beirut, the findings may not fully reflect the situation in other Lebanese hospitals or in different healthcare systems across the region. The cross-sectional design also provides only a snapshot in time and does not allow assessment of how knowledge and attitudes may change following education or clinical experience. Because the questionnaire was self-administered, some responses could have been affected by over- or under-reporting, or by misinterpretation of items, even though a validated tool was used. The study sample was dominated by nurses, with fewer physicians and pharmacists, which may have influenced subgroup comparisons. In addition, certain groups, such as pediatric healthcare providers, were not included, and the tool measured knowledge and attitudes rather than actual clinical practice, which may not always align. Finally, the modest sample size for some subgroups may have limited the statistical power to detect meaningful differences.

## Conclusion

The study results reflect the need for structured educational programs integrated into the teaching curricula on pain management based on the mean score of correct answers and the adequacy benchmark of ≥ 80 % achieved only by 5.6% of the total sample. The low mean scores of nurses and pharmacist reflect the need for educational and practical workshops to increase the interdisciplinary collaboration between HCWs in pain management. Overall, pain management in Middle East region as our study mirror with national and regional study results, requires change and improvement in pain management education for better patient pain control.

The authors are thankful to all who contributed to this article, including the participants who generously shared their time and experiences, and the hospital where the survey was conducted. Your support and cooperation were invaluable to the completion of this work.

## Data Availability

All data produced in the present work are contained in the manuscript

## Funding

The trial did not receive any funding from any source.

## Competing interests

The authors declare no financial relationships with any organizations that might have an interest in the submitted work, and no other relationships or activities that could appear to have influenced the submitted work.

## Ethical approval

The Institutional Review Board (IRB) of the hospital approved the study before data collection started. The study purpose, harm, voluntary participation, confidentiality, anonymity and the need for any further clarifications on the study were stated in the survey front sheet where eligible participants will consent for approval to participate. The participants were coded and the data collection and analysis were accessed and done by the study research team only.

## Transparency

The lead author, as the guarantor of this manuscript, confirms that it provides an honest, accurate, and transparent account of the study being reported. All significant aspects of the study have been included, and any deviations from the original study plan or protocol (if applicable) have been fully explained.

## Dissemination to participants and related patient and public communities

The authors plan to disseminate the study results to all study participants as soon as the study is published. The findings will also be shared at the national level, including through appropriate healthcare networks and institutions, and will be presented at relevant national conferences to ensure broad accessibility and impact.

## Provenance and peer review

Not commissioned; externally peer reviewed.

## Notes

### Competing Interest Statement

The authors have declared no competing interest.

### Funding Statement

This study did not receive any funding

### Author Declarations

Health Ethics committee of Clemenceau Medical Center gave ethical approval for this work

